# Decreased inflammatory cytokine production of antigen-specific CD4^+^T cells in NMDA receptor encephalitis

**DOI:** 10.1101/2020.09.15.20193979

**Authors:** Le-Minh Dao, Marie-Luise Machule, Petra Bacher, Julius Hoffmann, Lam-Thanh Ly, Florian Wegner, Alexander Scheffold, Harald Prüss

## Abstract

Anti-N-methyl-D-aspartate-receptor (NMDAR) encephalitis is the most common autoimmune encephalitis with psychosis, amnesia, seizures and dyskinesias. The disease is mediated by pathogenic autoantibodies against the NR1 subunit that disrupt NMDAR function. Antibody infusion into mouse brains can recapitulate encephalitis symptoms, while active immunization resulted also in strong T cell infiltration into the hippocampus. However, whether T cells react against NMDAR and their specific contribution to disease development are poorly understood. Here we characterized the ex vivo frequency and phenotype of circulating CD4^+^ T helper (T_H_) cells reactive to NR1 protein using antigen-reactive T cell enrichment (ARTE) in 24 patients with NMDAR encephalitis, 13 patients with LGI1 encephalitis and 51 matched controls. Unexpectedly, patients with NMDAR encephalitis had lower frequencies of CD154- expressing NR1-reactive T_H_ cells than healthy controls and produced significantly less inflammatory cytokines. No difference was seen in T cells reactive to the synaptic target LGI1 (Leucine-rich glioma-inactivated 1), ubiquitous *Candida* antigens or neoantigens, suggesting that the findings are disease-specific and not related to therapeutic immunosuppression. Also, patients with LGI1 encephalitis showed unaltered numbers of LGI1 antigen-reactive T cells. The data reveal disease-specific functional alterations of circulating NMDAR-reactive T_H_ cells in patients with NMDAR encephalitis and challenge the idea that increased pro-inflammatory NMDAR-reactive T cells contribute to disease pathogenesis.

## Introduction

Autoantibodies targeting the NR1 subunit of the N-methyl-D-aspartate-receptor (NMDAR) are the hallmark of NMDAR encephalitis (NMDAR-E)(Dalmau, Tüzün et al. 2007). The antibodies are directly functional, mainly by internalization of NMDAR and subsequent disruption of glutamatergic transmission(Planaguma, Leypoldt et al. 2015; Kreye, Wenke et al. 2016). Presence of the antibodies in the cerebrospinal fluid (CSF) results in clinical symptoms ranging from psychosis to amnesia to autonomic dysfunction(Dalmau, Armangue et al. 2019). The increasing knowledge on these antibodies showed differences to other autoimmune diseases. For example, NR1 antibodies in patients’ CSF had only few somatic hypermutations or were even unmutated, suggesting that T cell help might not be required for antibody development(Wenke, Kreye et al. 2019). Such germline antibodies were nonetheless pathogenic in established *in vitro* assays. On the other hand, T helper (T_H_) cells typically potentiate antibody secreting cell (ASC)-mediated antibody responses, and active immunization of mice with conformationally stabilized NMDAR protein resulted in profound infiltration of the hippocampus with not only ASC, but also CD4^+^ T cells(Jones, Tovar et al. 2019). CSF cytokine/chemokine signatures, such as increased interleukin-17, interleukin-6 or CXCL10 levels, also suggest a contribution of T cells in NMDAR-E(Byun, Lee et al. 2016; Liba, Kayserova et al. 2016), and brain biopsies or autopsies demonstrated infiltrates of CD4^+^ cells as well(Martinez-Hernandez, Horvath et al. 2011; Bien, Vincent et al. 2012). However, their antigen specificities are unknown. Until now, the characteristics of NR1-specific T_H_ cells in patients with NMDAR-E are poorly described. Using antigen-reactive T cell enrichment (ARTE)(Bacher, Schink et al. 2013), we therefore quantified and characterized the rare CD4^+^ T_H_ cells reactive to NR1 protein. Results provide insight into the immunological cascades leading to NMDAR antibody development and disease pathogenesis, might later help to explain the highly variable clinical spectrum in patients despite similar antibody titers, and could potentially lead to additional therapeutic approaches beyond antibody-directed therapies.

## Methods

### Cohorts

All clinical investigations were conducted according to Declaration of Helsinki principles. Written informed consent was received from participants at the Centre for Autoimmune Encephalitis, Department of Neurology, Charité – Universitätsmedizin Berlin, prior to inclusion into the study and analyses were approved by the Charité Institutional Review Board. Participants included (1) 24 patients with NMDAR-E, (2) 33 age- and sex-matched controls for NMDAR-E, (3) 13 patients with LGI1 (Leucine-rich glioma-inactivated 1) encephalitis and (4) 18 age- and sex-matched controls for LGI1-E. Encephalitis patients fulfilled diagnostic criteria, i.e. the characteristic clinical picture together with high-titer NMDAR/LGI1 autoantibodies(Graus, Titulaer et al. 2016).

### Proteins for T cell stimulation

We used commercially available NR1 and LGI1 proteins (MyBiosource, 964741 and 1378533). We further generated fragments of the extracellular part of the NMDAR NR1 subunit containing the aminoterminal domain (ATD) or the S1 and S2 domains fused together (S1S2) (Fig. 1A). Proteins were expressed as GST-fusion proteins in *E.coli* and purified via glutathione-Sepharose^TM^ 4B (GE Healthcare, 52-2303-00). All four proteins showed high purity in coomassie staining after gel electrophoresis (Fig. 1B).

**Figure 1.**
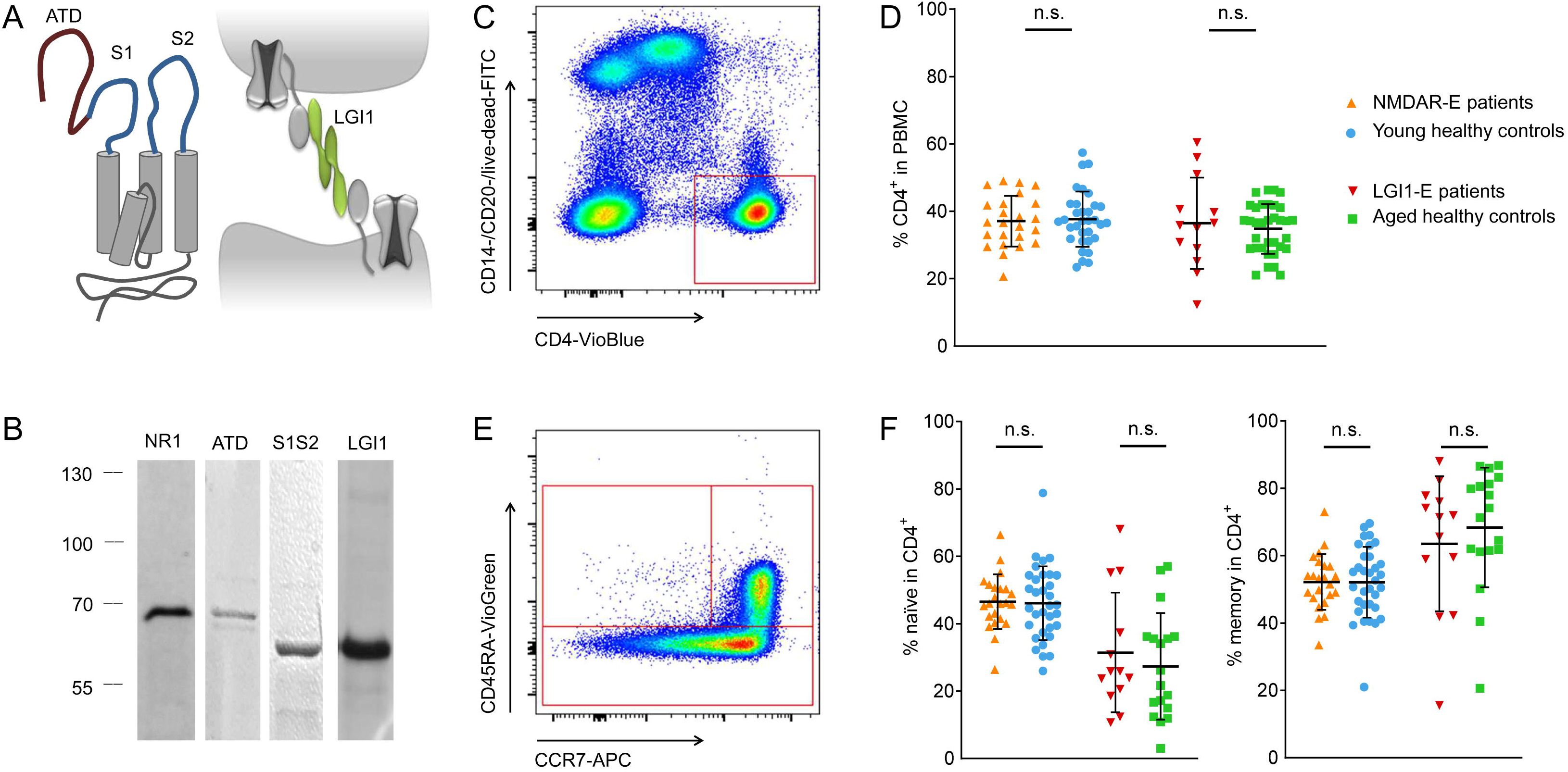
Antigens, flow cytometry gating strategy and CD4^+^ T cell frequencies. ATD (aminoterminal domain), S1 and S2 are extracellular domains of the NR1 subunit of the NMDA receptor, while LGI1 is part of a trans-synaptic protein complex that regulates voltage-gated potassium channels (A). All four proteins used for antigen stimulation in this study showed high purity in coomassie staining after gel electrophoresis (B; ATD + GST tag = 68 kDa, S1S2 + GST tag = 57 kDa, LGI1-his-myc = 64 kDa, NR1-his-myc = 68 kDa). No differences in CD4^+^ T cell frequencies within PBMCs were observed in all cohorts (C-D). CD4^+^ T cells were gated for CCR7+ CD45RA^-^ to measure dividing memory and naïve CD4^+^ T cells (E). NMDAR-E and LGI1-E patients showed similar distributions of naïve and memory CD4^+^ T cells compared to their respective controls. As expected, older subjects (LGI1-E patients and their controls) had more memory and less naïve T cells than younger subjects (F).

### Antigen-reactive T cell enrichment (ARTE)

Enrichment of NMDAR/LGI1 antigen-specific T cells followed established protocols(Bacher, Schink et al. 2013). In brief, peripheral blood mononuclear cells (PBMC) were isolated from BD Vacutainer® CPT^tm^ venous blood samples, washed with PBS, quantified and resuspended in RPMI with 5% human AB serum (Sigma-Aldrich, H4522). 1×10^7^ cells in 1 ml were incubated in 12-well plates for 12 hours at 37°C with 5% CO_2_. Each well was stimulated with 10 μg autoantigen, together with negative and positive controls for 7 hours at 37°C. One sample per participant remained unstimulated to determine the ratio of CD4^+^ T cells (original fraction, ORI) from PBMCs to allow calculation of the CD154^+^ cell frequency for each sample.

Internalization of CD 154 on activated T cells was inhibited during stimulation by blocking CD40 on antigen presenting cells with 1 μg/ml CD40 antibodies (Miltenyi, 130-094-133, 100 μg/ml). After 5 hours, 1 μg/mL Brefeldin A (Sigma-Aldrich, B6542) was added to inhibit the secretion of activation markers. *C. albicans* lysate (Stallergenes Greer, XPM15D3A5) at 20 μg/ml served as positive control. Negative controls were added at equal volumes depending on the autoantigen (PBS for ATD and S1S2; 10 mM Tris-HCL [with 1 mM EDTA and 20% glycerol] for commercial LGI1 and NR1 proteins). Keyhole limpet haemocyanin (KLH) at 200 μg/ml served as neoantigen control (Stellar Biotechnologies, KLH-05NR), confirming that the assay detects all antigen-activated cells including from the naïve repertoire. NMDAR/LGI1-reactive T cells were isolated using the anti-CD 154 Microbead Kit human (Miltenyi Biotec, 130-092-658) using MS columns (Miltenyi, 130-042-201).

### Flow cytometry

Cells were stained against CD4 (Miltenyi, 130-113-219, Vioblue, human, 1:50; 130-096-652, APC-Vio770, human, 1:11), CD14 (Miltenyi, 130-098-063, FITC, human, 1:11), CD20 (Miltenyi, 130-091-108, FITC, human, 1:11), CD45RA (Miltenyi, 130-096-921, CD45RA- Vio®Green, human, 1:11), CD45RO (Miltenyi, 130-099-044, VioBlue, human, 1:11) and CCR7 (Miltenyi, 130-108-286, APC, human, 1:11). Intracellular stainings included IFN- gamma (Miltenyi, 130-109-231, PE, human, 1:11; Biolegend, 502526, PerCP-Cy5.51, human, 1:100), TNF-alpha (Miltenyi, 130-096-755, PE-Vio®770, human 1:11) and CD154 (Miltenyi, 130-109-474, PerCP-Vio700, human, 1:11; Miltenyi, 130-092-289, PE, human, 1:11). Samples were measured with flow cytometry (BD FACSCanto II, BD, 338962) and analyzed with FlowJo® (BD, FlowJo™ v10.5.3) (Fig. 1C, E).

### Statistical analysis

GraphPad Prism software (GraphPad Software, Version 7.5) was used for statistical analysis. For all measurements, background activity (CD154^+^ T cells in PBS/buffer control samples) was subtracted. The Shapiro-Wilk and D’Agostino Pearson tests were used to test for normal distribution of CD154^+^ T cell frequencies. As normal distribution was found, all 4 cohorts were analyzed with one-way ANOVA and corrected for multiple comparisons with Tukey tests. If two groups were compared, two-sided t-tests were used for independent samples. In all graphs, average values are shown with standard deviations. The significance level was defined as P<0.05.

## Results

### Study participants

Study cohorts included patients with NMDAR-E (n=24), patients with LGI1-E encephalitis (LGI1-E) (n=13) and respective healthy controls. As NMDAR-E is a disease of predominantly young female patients and LGI1-E a disease of aged male patients, two different age- and sex-matched groups of unaffected controls were recruited (n=33 and n=18, respectively). The mean age was 31.9 ± 11.9 years [± SD] in NMDAR-E patients (3 M, 21 F), 34.4 ± 13.5 years in their controls (10 M, 23 F), 68.7 ± 13.1 years in LGI1-E patients (8 M, 5 F) and 69.2 ± 10.5 years in their controls (13 M, 5 F).

### NMDAR-E patients had reduced frequencies of NR1-reactive T cells

All patients and controls had similar frequencies of total CD4^+^ T cells in PBMCs (Fig. 1C-D). As expected, elder cohorts had more memory and less naïve CD4^+^ T cells than younger subjects, equally for patients and healthy controls (Fig. 1E-F).

After incubation of PBMCs with commercially available full-length extracellular NR1 protein in a small pilot cohort, antigen-reactive CD 154-expressing CD4^+^ cells were isolated via magnetic cell separation and determined with flow cytometry (Fig. 2A). As a quality control, duplicate measurements of *C. albicans* stimulation from eight patients on the same day showed high correlation of CD154^+^ cell frequencies (Fig. 2B). T cells reactive to NR1 protein were significantly reduced in patients with NMDAR-E compared to controls (Fig. 2C). The levels of inflammatory cytokines in CD154^+^ (Fig. 2D) and CD4^+^ cells (Fig. 2E) were not significantly different between groups.

**Figure 2.**
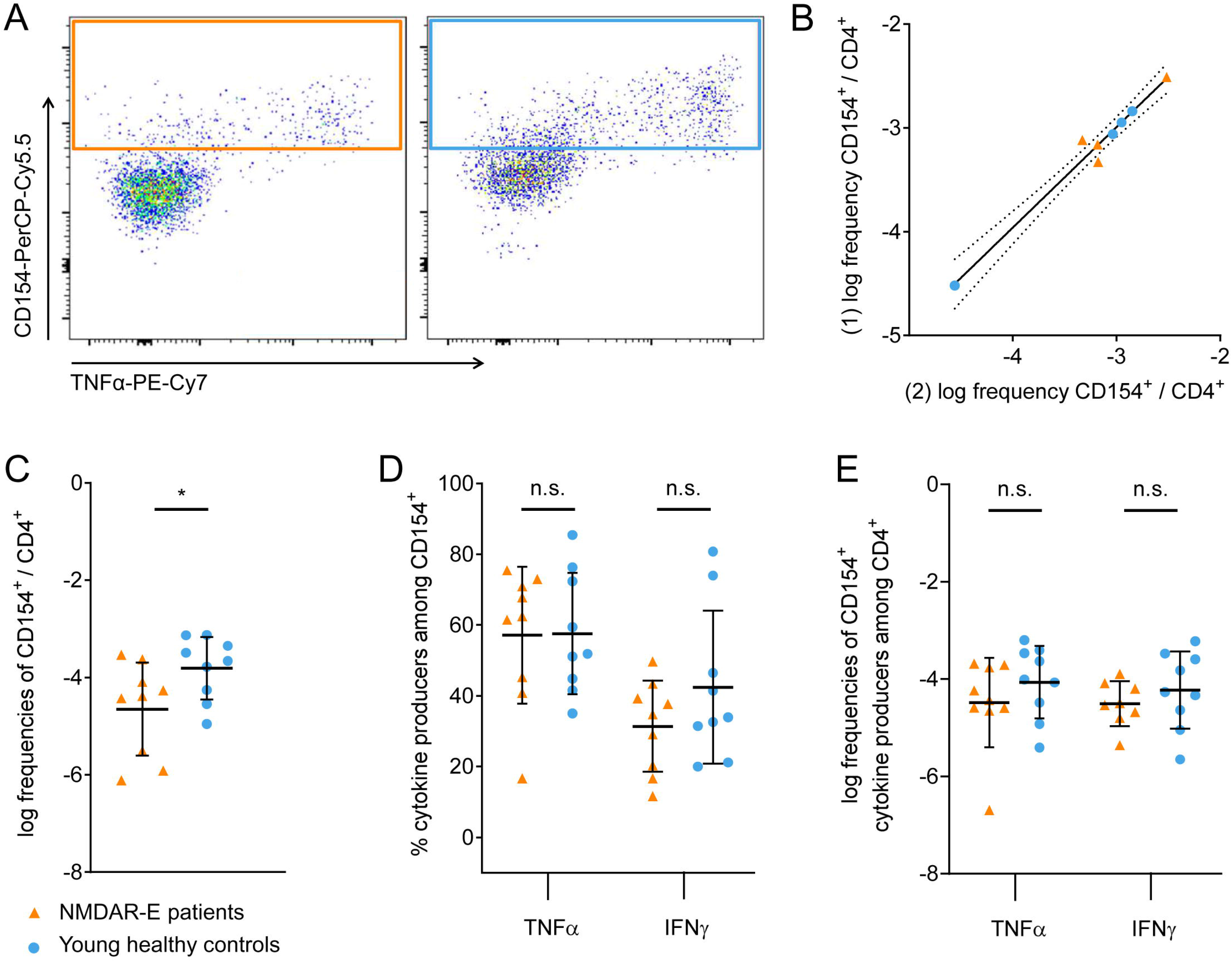
Antigen-specific CD154^+^ CD4^+^ T cells in NMDAR-E. Frequencies of antigen-reactive*** CD 154-expressing T cells (exemplary gating in A) were decreased in NMDAR-E patients compared to controls after stimulation with recombinant full-length extracellular NR1 protein (C). Percentage of TNFα and IFNγ-expressing antigen-specific CD4^+^ T cells was not different between NMDAR-E patients and controls after stimulation with full-length NR1 (D-E). Biological replicates of *C. albicans* stimulation from PBMCs of the same patient showed robust correlation of CD154^+^ T cell frequencies in separate measurements, indicating low variability in the assay system (B).

### NMDAR-E patients had reduced frequencies of CD154^+^ T cells reactive to the extracellular S1S2 domain of the NMDAR

Following the unexpected observation that NR1-reactive CD154-expressing CD4^+^ T cells were reduced in NMDAR-E, we next examined the antigen reactivity to smaller regions of the extracellular NR1 domains (Fig. 1A-B) in a larger cohort of patients. We selected the aminoterminal domain (ATD) – the main target epitope of pathogenic autoantibodies in NMDAR-E – and the S1+S2 domains, which are the extracellular parts of the NMDAR NR1 subunit. T cells reactive to the S1S2 domain and to the combination of ATD and S1S2 were significantly reduced in patients with NMDAR-E compared to controls (Fig. 3A). Although not reaching statistical significance, a similar trend was detected after stimulation with ATD alone. In contrast, patients with NMDAR-E and healthy controls showed no difference in T cells responding to the synaptic protein LGI1 or the neoantigen keyhole limpet haemocyanin (KLH) (Fig. 3A). As expected, the absolute frequency of KLH-specific cells was lower compared to NR1-specific cells(Bacher and Scheffold 2013).

**Figure 3.**
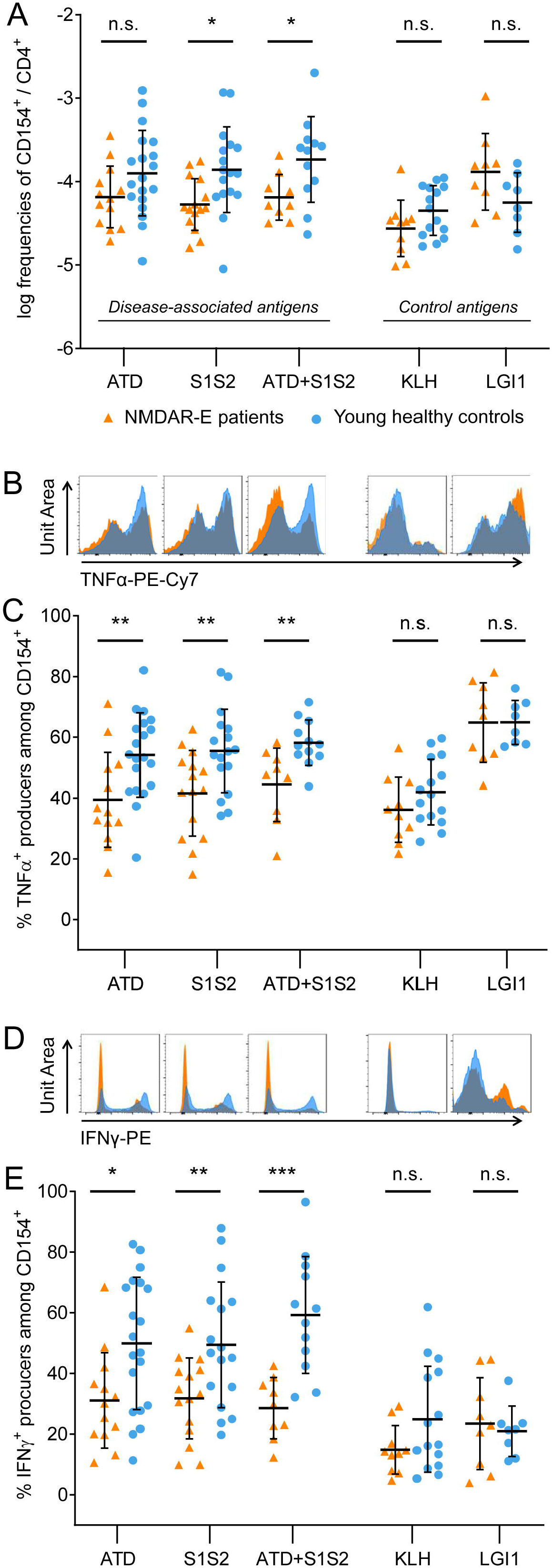
T cell reactivity to fragments of extracellular NR1 and reduced inflammatory cytokine production of CD154^+^ CD4^+^ T cells in NMDAR-E patients. Using smaller regions of the extracellular NR1 domain in an expanded cohort of patients with NMDAR-E, frequencies of antigen-reactive CD154-expressing T cells were significantly decreased compared to controls after stimulation with S1S2 or the combination of ATD and S1S2, but did not reach statistical significance with ATD alone (A, left). Numbers of antigen-reactive T cells to the neoantigen KLH or the synaptic protein LGI1 were not different in NMDAR-E patients (A, right; logarithmic frequencies within CD4^+^ T cells). Antigen-specific CD154CD4^+^ T cells from NMDAR-E patients showed significantly lower percentages of TNFα-expressing cells after stimulation with ATD, S1S2 or ATD+S1S2 compared to healthy controls, but not after stimulation with KLH or LGI1 control proteins (B- C; histograms display pooled group data). Similarly, antigen-specific T cells from NMDAR-E patients showed significantly lower percentages of IFN -expressing cells after stimulation with ATD, S1S2 or ATD+S1S2 compared to controls, but not after KLH or LGI1 stimulation (D-E).

### Reduced production of inflammatory cytokines in NMDAR-E CD154^+^ T cells

We next determined whether antigen-activated T cells from patients with NMDAR-E differed in the expression levels of inflammatory cytokines. CD4^+^ T cells showed significantly reduced frequencies of TNFα^+^ after incubation with ATD, S1S2 proteins and the combination of both stimulations (Fig. 3B-C). No difference was observed in the TNFα^+^ production of CD4^+^ T cells from NMDAR-E patients reactive to KLH and LGI1 (Fig. 3B-C).

A qualitatively similar picture was seen when comparing the expression of IFNγ^+^ CD154^+^ T cells between patients with NMDAR-E and controls. ATD, S1S2 and the combination of both proteins elicited a significantly smaller IFNγ response in patients compared to healthy subjects (Fig. 3D-E), while the frequencies of IFNγ^+^ cells after KLH and LGI1 incubation were equal between both groups (Fig. 3D-E).

### No changes of antigen-reactive T cells in LGI1-E patients

To determine whether reduced frequencies of antigen-reactive T cells are a common phenotype across autoimmune encephalitides, we expanded our study to LGI1-E, the second most common form after NMDAR-E. However, patients with LGI1-E did not show an altered frequency of T cells reactive to full-length LGI1 (Fig. 4A), suggesting disease-specific effects in NMDAR-E patients. Also in contrast to NMDAR-E, TNFα- and IFNγ-positive T cells after stimulation with the disease-defining antigen LGI1 were not altered in LGI1-E patients and controls (Fig. 4B-C).

**Figure 4.**
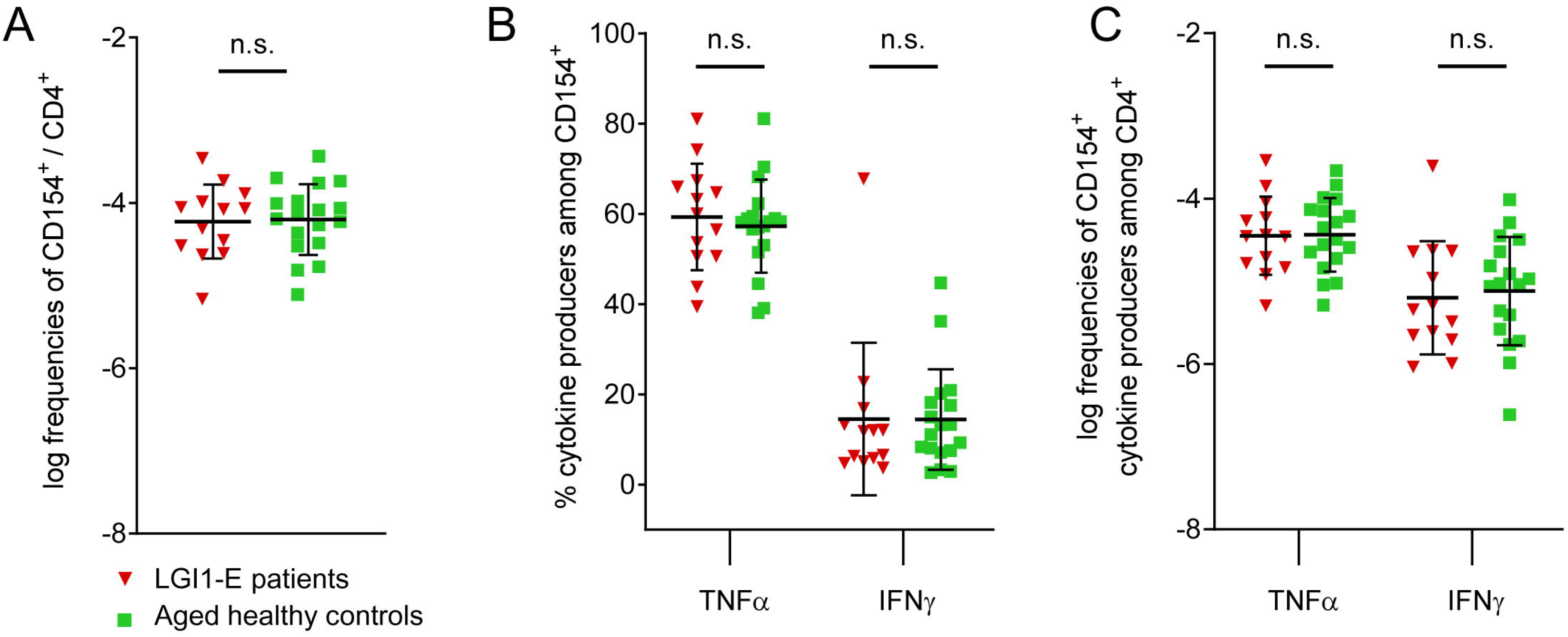
Antigen-specific CD154^+^ CD4^+^ T cells in LGI1-E. In contrast to NMDAR-E, patients with LGI1-E showed similar frequencies of CD154^+^ T cells after stimulation with its disease- specific antigen LGI1 compared to controls (A). LGI1-E patients did not show differences in TNFα or IFNγ production after stimulation with LGI1 (B-C).

### Diminished antigen-specific CD154^+^ T cell frequencies are not related to immunosuppression

To exclude that the reduction of NR 1-activated T cells in NMDAR-E patients is related to previous treatments with immunosuppression, we next determined the T cell activation by the ubiquitously available *Candida albicans* antigen to which typically all people have strong T cell reactivity(Bacher and Scheffold 2013; Koehler, Cornely et al. 2018). No differences were observed in the frequency of Candida-specific CD 154-expressing T cells of patients with NMDAR-E, LGI1-E and both healthy control groups (Fig. 5A). Similarly, the production of TNFα and IFNγ were not different between all four groups (Fig. 5B) suggesting unimpaired immune function in patients.

**Figure 5.**
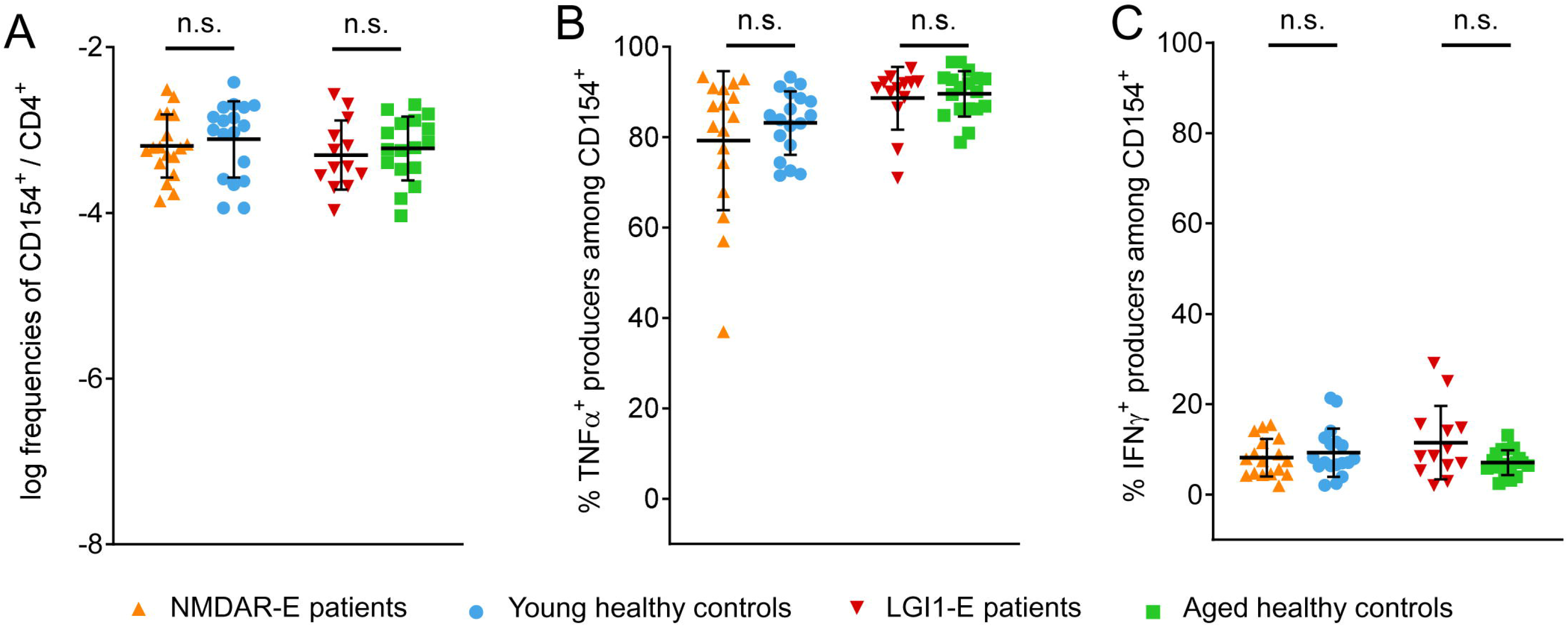
Equal numbers of T cells reactive to *Candida albicans*. All patients and controls showed similar frequencies of antigen-specific CD154^+^ CD4^+^ T cells after stimulation with *C. albicans* lysate, indicating that observed differences after NMDAR stimulation were not caused by general immunosuppression (A). Similarly, TNFα and IFNγ responses were not different between groups (B-C).

## Discussion

The present study is to our knowledge the first direct ex vivo quantitative and qualitative analysis of circulating autoreactive T cells involved in NMDA receptor encephalitis. Our data reveal the unexpected finding that NR1-reactive CD4^+^ T_H_ cells are not elevated but rather reduced in patients with NMDAR-E and that these cells produce less inflammatory cytokines. No difference was seen in T_H_ cells reactive to the synaptic target LGI1, ubiquitously available *Candida* antigens or neoantigens when comparing NMDAR-E patients with controls. In contrast, patients with LGI1-E did not have reduced inflammatory cytokines and T_H_ cell frequencies against their disease-defining antigen LGI1. The data collectively suggest that the reduced T_H_ cell numbers in NMDAR-E are disease-specific.

Based on our findings, reduced NR1-specific T_H_ cell numbers are not related to therapeutic immunosuppression. Although most patients with NMDAR-E in this cohort received first- and second-line immunotherapy, the T_H_ cell immune response against the control antigens was unaltered, including reactivity to *Candida* antigens, the neoantigen KLH and the synaptic protein LGI1. The frequencies of autoantigen-specific T_H_ cells observed here were in the range of previous studies, i.e. 1 in 10^4^-10^6^ T cells(Bacher and Scheffold 2013).

For the selection and production of highly specific and highly affine antibodies, T_H_ cells generally mediate the T cell-dependent affinity maturation and enhance antibody responses. Given the strong antibody-mediated etiology in NMDAR-E, we therefore expected rather increased numbers of NR1-specific T_H_ cells and increased expression of inflammatory cytokines as shown in comparable diseases. For example, in the related antibody-defined CNS disease neuromyelitis optica (NMO), T_H_ cells against the underlying protein aquaporin-4 (AQP4) were clonally expanded and demonstrated greater proliferation to AQP4 than T cells from healthy controls(Varrin-Doyer, Spencer et al. 2012). However, these data were generated following long-term *in vitro* culture which may select individual T cell clones and significantly alter their phenotypic and functional characteristics. Experimental findings involving AQP4 knockout mice rather suggested that the T cell response to AQP4 is normally regulated by stringent mechanisms of central and peripheral tolerance, indicating dysfunctional thymic deletion in NMO(Sagan, Winger et al. 2016).

However, contrary to our hypothesis, we found a reduction of NR1-reactive T_H_ cells and markedly decreased inflammatory cytokines in these cells in patients with NMDAR-E. It seems possible that T_H_ cells expressing CD154 (the very important B cell helper molecule) might not be required for NR1 antibody generation, thus extrafollicular B cell activation may occur independently of T cell help, e.g. directly in the brain or in some patients in NMDAR-expressing tumor cells in ovarian teratomas, a common association in women with NMDAR- E(Day, Laiq et al. 2014). In fact, the finding that pathogenic patient-derived NMDAR antibodies had only few or even lacked somatic hypermutations(Wenke, Kreye et al. 2019) is consistent with a non-germinal center origin, not seen in monoclonal antibodies of patients with LGI1-E(Kornau, Kreye et al. 2020). Pathogenic unmutated antibodies are also in striking contrast to NMO, where AQP4 antibodies lost any detectable binding to AQP4 when they were reverted back to their unmutated germline precursors, indicating the need for somatic hypermutations (and thus involvement of T_H_ cells) to cause disease(Cotzomi, Stathopoulos et al. 2019).

A less likely possibility for the reduced NR1-specific T_H_ cell numbers could be the recruitment to germinal centers in the lymphatic organs, to germinal center-like structures in tumors(Day, Laiq et al. 2014) and brain, or to the brain parenchyma, thus not being detectable in the blood. In the murine model of NMDAR-E following active immunization, CD4^+^ T cells were indeed the predominant class of immune cells in the hippocampus at three weeks(Jones, Tovar et al. 2019). The limited availability of biopsy and autopsy samples will make it very difficult to confirm this hypothesis in humans. T cell receptor sequencing to compare clones between CSF and blood or repeated analyses of the T cell frequencies in the same patients during the disease course might partially help as normalization of cell numbers may occur with time. Theoretically, overstimulation/exhaustion of NR1-specific T_H_ cells or expansion of regulatory T_H_ cells (T_reg_) might also lead to reduced numbers of NR1-specific T_H_ cells. It would be difficult to predict, however, in which way T_reg_ were stimulated in NMDAR-E, keeping in mind that it has not been possible yet to isolate and functionally characterize autoantigen-specific T_reg_ in humans.

Our study further suggests that a similar reduction of T_H_ cell reactivity toward the disease-defining antigen is not a universal finding in the still expanding types of autoimmune encephalitis. Patients with LGI1-E in the present study did not show altered frequencies of LGI1-reactive T_H_ cells, and it will be interesting to see the T cell phenotype of additional encephalitides in the future, such as GABA_a_R, IgLON5, mGluR5 or Caspr2 encephalitis. It is possible that immuno-genetic factors can influence the profiles of immune cell frequencies and distribution. Interestingly, while LGI1-E was strongly linked to the HLA-DR7 haplotype(van Sonderen, Roelen et al. 2017; Mueller, Farber et al. 2018), NMDAR-E showed an association with HLA class II allele DRB1*16:02 in one study, which has previously been found in autoimmune diseases predominantly mediated by autoantibodies(Shu, Qiu et al. 2019).

Taken together, we unexpectedly observed a reduced number and profound disease-specific functional alterations of NR1-reactive T_H_ cells in patients with NMDAR-E that were not related to immunotherapies, challenging the idea that increased pro-inflammatory NMDAR- reactive T_H_ cells contribute to disease pathogenesis. However, the significant modulation of the autoantigen-specific T cell response clearly reveals an autoimmune reaction in these patients. One may hypothesize that, following an initial priming phase, NR1 autoantibody production might not depend on continuous T cell help. Follow-up studies including T cell receptor sequencing as well as generation and functional characterization of T cell lines are needed to further characterize the dysfunctional cytokine response.

## Data Availability

all raw data are available on our local Charite server

